# Anxiety Levels among Healthcare Professionals during Covid-19 Pandemic: A Multifactorial Study

**DOI:** 10.1101/2020.10.14.20212167

**Authors:** Arif Malik, Muhammad Mansoor Hafeez, Sulayman Waquar, Muhammad Asim Rana, Rabail Alam

## Abstract

The current study focuses on psychological stress level among doctors, estimated by calculating anxiety score. For the assessment of anxiety levels, the GAD-7 scale was used. Chi-Square test and Odd ratios were calculated among the exposed and not exposed groups involved in the management of COVID-19 patients. Results revealed increased anxiety levels in the exposed group. Besides, the availability of personal protective equipment’s and stress from the family to quit the job were the substantial contributing factors that increased anxiety. Based on the results, it is proposed that the concern administrative authorities should consider these findings to facilitate medical healthcare professionals.

## Introduction

Anxiety has been identified as one of the top-ranked mental issues among health care professionals that can dynamically influence individual performances. Different external factors like working hours at the hospital, personal relationships, financial problems and certain other extraneous factors including medical illness of family members can contribute to elevated stress levels (Koay et al., 2020). It is pertinent to mention here that anxiety and depression levels generally prevailing among the population worldwide (Fonagy et al., 2016). Such non-communicable disorders could potentially lead to disability (Murray et al., 1996). More specifically, in developing countries like Pakistan, psychiatric disorders in combination with numerous infectious diseases and malnutrition can further aggravate the problem. On a gender basis, women are more prone to this disorder compared with male counterparts (Mirza et al., 2004). Keeping in view the available literature, the present study investigated anxiety levels among healthcare professionals during COVID-19 pandemic that added extra stress among the medical community, especially frontline healthcare professionals. So far, the COVID-19 infection has caused casualties including 278 doctors that were confirmed but more than reported deaths of doctors are expected (Ing et al., 2020). The novel coronavirus disease (COVID-19 or 2019-nCoV) was first identified in December 2019 in China and the infection spread to 167 countries within a few months. Apart from the rapid rise in death tolls, COVID-19 infection rates caused added stress among the general population that ultimately affected front line doctors working in hospitals (Montemurro et al., 2020). World Health Organization (WHO) declared it as a pandemic/global emergency in March 2020 due to an increasing number of cases and deaths around the globe. According to WHO, there were 3.7 million confirmed cases with a death toll of 1 million (WHO COVID-19 Dashboard., 2020). Outbreaks of COVID-19 have imposed critical impacts on the mental health and anxiety-related concerns among healthcare professionals (Ahmed et al., 2020).

It should be noted that a large number of doctors and nurses have got infected and even lost their lives in this pandemic. Major existing issues were due to increased workload and intermittent lack of protective equipment (Magnani et al., 2020). Researchers integrated a significant impact of the pandemic with psychological and emotional behaviors of healthcare professionals (Spinelli et al., 2020). Unfortunately, the major focus of allied healthcare professionals was to identify and to elucidate the pathophysiology of COVID-19 diseases. Somehow, the mental health condition of those who were actively involved in treating a mass of the population was neglected or very little focused. It this extremely unavoidable situation, a very important aspect of anxiety levels among health professionals therefore remained unnoticed. In the present study, we investigated the prevalence of anxiety levels and associated contributing factors among health care physicians during COVID-19 pandemic.

## Materials and Methods

### Experiment details

A total of 156 doctors were recruited to study the mental stress and anxiety levels during a cross-sectional comparative study from April to June (2020). They were further divided into two groups. Participants of group A were never directly exposed while those of group B were exposed to the treatment of COVID-19 patients.

### Assessment Questionnaire and GAD-7 Scoring

Questionnaires were distributed among the participants from both groups. Responses were collected from different hospitals of Lahore (31.5204° N; 74.3587° E) Punjab, Pakistan. Doctors having already some mental illness or on anti-psychotics were excluded from the study. For the assessment of anxiety GAD-7 scoring system was used and outcomes were assessed (Spitzer et al., 2006). Scores obtained in between 0-21 was encoded as follows: Scores of 5-9 for mild anxiety, 10-14 moderate anxiety, and ≥ 15 severe anxiety. Apart from the GAD-7 scoring two other factors a) Family stress to quit the job, b) Lack of adequate supply of personal protective equipment’s (PPE’s) were also assessed which were specifically relevant to group B.

### Statistical Analyses

The data were statistically analyzed with the help of a Chi-square test using SPSS software (version 20). In addition, the p-values of less than 95 % confidence index (*p*<0.05) remained statistically significant.

## Results

### Anxiety levels among doctors

The study assessed different variables in the identification of anxiety levels within the doctors. Male to female ratio in group A was 36:25, whereas, in group B, it was 64:31. Chi-square analysis depicted no significant difference in anxiety levels of two groups based on gender (p=0.187). By contrast, significantly increased levels of anxiety were recorded among members of group B (p=0.005) that are directly in contact with COVID-19 patients (Table-1).

**Table-1:** Changes in Anxiety Levels in Doctors Calculated by Using GAD-7 scoring.

|  |  | Doctors |  | Total | *p*-value |
| --- | --- | --- | --- | --- | --- |
|  |  | Group A | Group B |  |  |
| **Gender** | Male | 36 (36.0%) | 64 (64.0%) | 100 (100.0%) | 0.187 |
|  | Female | 25 (44.6%) | 31 (55.4%) | 56 (100.0%) |  |
| **Anxiety Score** | No Anxiety | 38 (52.8%) | 34 (47.2%) | 72 (100.0%) | 0.005 |
|  | Mild Anxiety | 6 (20.0%) | 24 (80.0%) | 30 (100.0%) |  |
|  | Moderate Anxiety | 13 (37.1%) | 22 (62.9%) | 35 (100.0%) |  |
|  | Severe Anxiety | 4 (21.1%) | 15 (78.9%) | 19(100.0%) |  |

Odds ratios were calculated by 2×2 cross-tabulation that explained the risks of anxiety in doctors were almost three times (OR=2.96) greater in Group B compared to Group A.

### Factors contributing to Aggravation of Anxiety Levels

Doctors included in the study further were inquired about the factors that contributed to the aggravation of their anxiety levels. Of these, two major contributing factors included stress from the family to quit the job and the availability of proper personal protective equipment’s (PPE’s). The findings indicated that there are 4.92 times and 1.80 times greater risk of anxiety in those who were facing stress because of inadequate supply of PPEs and from their families respectively (Table-2).

**Table 2:** Factors involved in the aggravation of anxiety levels among doctors.

| Doctors |  | Anxiety |  | Total | Odd Ratio (95% CI) |
| --- | --- | --- | --- | --- | --- |
|  |  | Yes | No |  |  |
| **Exposure** | Yes | 61 (64%) | 34 (36%) | 95 | 2.96<br>(1.52-5.77) |
|  | No | 23 (38%) | 38 (62%) | 61 |  |
| **Lack of PPE** | Yes | 28 (85%) | 5 (15%) | 33 | 4.92<br>(1.68-14.41) |
|  | No | 33 (37%) | 29 (53%) | 62 |  |
| **Family Stress** | Yes | 17 (74%) | 6 (26%) | 23 | 1.8<br>(0.63-5.12) |
|  | No | 44 (61%) | 28 (39%) | 72 |  |

## Discussion

The COVID-19 is the most devastating and challenging public health crises since the influenza pandemic in 1918. Global statistics show millions of cases and about half of million deaths by May 2020 (WHO, 2020). Our results indicated that professionals engaged in the treatment of COVID-19 patients were significantly influenced by anxiety. In agreement with our findings, it has been indicated that linear spread of the virus has undeniable effects over the population worldwide but doctors and nurses who are in direct contact with the affected individuals deal with constant stress and fear that drives them into an anxious state of mind (Shah et al., 2020). Significant efforts had been made worldwide in ruling out anxiety and anxiety-related issues from the health care professionals (Van Bavel et al., 2020). Fear of being a vector in affecting others i.e., (family, friends, relatives, neighbours etc.) due to the inadequate PPEs and appropriate preventive measures is an important social aspect that augments anxiety (Bauchner et al., 2020; Lam et al., 2020). The current study also investigated the involvement of family members and other associated factors to study changes in the magnitude of anxiety levels. Our results revealed that the lack of PPE’s, and stress from family to quit the job considerably increased the levels of anxiety among the healthcare professionals. Thus, it is the need of the hour that administrative departments and governing authorities should take positive steps in ruling out the concerns of anxiety and fears with which healthcare professionals and continuously engaged.

## Conclusions

From the results, it is evident that levels of anxiety among health care professionals are directly associated with i) direct exposure to COVID-19 infected patients ii) family support and iii) personal protective equipment (PPEs). It is, therefore, concluded that everyone has to play his role to cope with the situation like a pandemic. Modifiable aggravating factors for anxiety levels like proper provision of PPEs can be easily managed with the cooperation of hospital management and administrative officials, it may eradicate the fears of healthcare professionals to work in such a pandemic situation. Similarly, family support is also linked with the anxiety free and well protected environment of hospitals.

## Data Availability

I being the researcher have all data in the form of filled GAD-7 response sheets as well as in excel sheets

